# Circadian-related hypothalamic structure differs by chronotype in bipolar disorder

**DOI:** 10.64898/2026.01.27.26344976

**Authors:** Marlene Tahedl, Jonas Rohrer, Erich Seifritz, Daniel J. Smith, Philipp Homan

## Abstract

**Background:** Circadian rhythm disturbances represent a core feature of bipolar disorder (BD), with evening chronotype as a marker for poorer outcomes. We hypothesized that BD psychopathology combined with evening chronotype is associated with structural alerations in circadian-related hypothalamic regions – particularly the suprachiasmatic nucleus (SCN) – specific to BD relative to other psychiatric diagnoses.

**Methods:** We investigated structural neuroimaging data from the UK Biobank (113 BD, 205 major depressive disorder, 91 psychotic disorders, 199 healthy controls). The SCN-containing anterior-inferior hypothalamic subunit was segmented, central to circadian functional neuroanatomy. For each group, diagnosis × chronotype interactions on its volume were tested using analysis of variance, with post-hoc estimated marginal means and correction for multiple comparisons. Covariates included age, sex, handedness, and lithium use. Specificity was examined across four additional hypothalamic subunits.

**Results:** There was a diagnosis × chronotype interaction in the SCN-containing anterior-inferior hypothalamic subunit volume (*F*(6, 590) = 2.87, *p* =.009). This was driven by larger volumes in BD individuals with evening versus morning chronotype (*t* = 3.24, *p*_*FWER*_ =.004). No comparable results were found in other hypothalamic regions or diagnoses.

**Conclusions:** Hypothalamic structure differs by chronotype in BD, with chronotype related associations localized to an anterior-inferior hypothalamic region implicated in circadian regulation. These findings support chronotype as a biologically meaningful dimension of variation in BD and provide neuroanatomical evidence linking circadian preference to circadian relevant brain structure. Longitudinal and interventional studies will be important to clarify the temporal dynamics, underlying mechanisms, and potential clinical significance of these associations.

## Introduction

Bipolar disorder (BD) is a severe and highly variable psychiatric condition associated with substantial morbidity, elevated suicide risk, and marked functional impairment (1). Despite major advances in pharmacological and psychosocial treatment, outcomes remain variable, highlighting the need for biologically informed stratification approaches (2,3).

Disruptions of circadian regulation are a core feature of BD and have been implicated in its pathophysiology across multiple levels of analysis. Notably, circadian-related pathophysiology has been argued to be particularly prominent in BD as opposed to other major psychiatric disorders including major depressive disorder (MDD, (4,5)). Clinically, hallmark symptoms of BD, including fluctuations in mood, energy, activity, appetite, and sleep, are tightly coupled to circadian rhythms (6). Sleep wake dysregulation is particularly prominent, with reduced need for sleep during manic episodes and prolonged sleep or phase delays during depressive episodes. Converging evidence from neuroimaging, electrophysiological, and molecular studies further implicates circadian related neural systems in BD (7), motivating growing interest in circadian based therapeutic interventions (8,9).

Interventions targeting circadian regulation, most commonly bright light therapy (BLT), have shown therapeutic benefits in BD (10), but also in other psychiatric conditions including MDD or seasonal affective disorder (SAD) (11). However, emerging evidence indicates distinct mechanisms of BLT in BD as compared to MDD or seasonal affective disorder (SAD). In BD, BLT primarily induces circadian normalization, whereas in MDD/SAD it enhances mood via non-circadian pathways, such as serotonergic modulation and alertness (12–15). These distinct effects arise from diverging neural pathways. The circadian pathway propagates light information via intrinsically photosensitive retinal ganglion cells (ipRGCs, especially M1 subtype) through the retinohypothalamic tract (RHT) to the central circadian pacemaker, the suprachiasmatic nucleus (SCN), with indirect projections to the melatonin-producing pineal gland modulating phase advances or delays. In contrast, the mood-enhancement pathway in MDD propagates via ipRGCs (especially M4 subtype) through the ventral lateral geniculate nucleus and lateral habenula (inhibition) to the limbic system (16–20). These pathways are supported by the persistence of non-circadian mood enhancement after SCN ablation (14,21,22). While low-intensity (∼120–500 lux) has been reported to suffice for circadian entrainment, mood benefits may require higher intensities (>5,000 lux) (23,24).

However, independent of the neural / therapeutic pathways, not all individuals appear to benefit equally from BLT (25–27). Identifying patient characteristics associated with differential responsiveness remains an important open question. Chronotype, defined as an individual’s preferred timing of sleep and activity, is a stable and partly heritable dimension of circadian biology (28). Evening chronotype has been associated with greater symptom severity in BD (29) and other psychiatric disorders, as well as with altered sleep and activity patterns (30–32). Notably, there is evidence suggesting that evening chronotype individuals with BD may show enhanced responsiveness to BLT (33). Similar observations have been reported for MDD and various other conditions (34,35). While these findings point to chronotype as a potentially relevant source of clinical variability, the differential neural pathways described above suggest that chronotype related effects may differ biologically between BD and other conditions.

One possibility is that the differential chronotype-related differences in BLT-responsiveness in BD as opposed to e.g. MDD are associated with an enhancement of circadian rhythm disruption. Since profound desynchronization is a pathological hallmark feature of BD, but not of MDD, we therefore asked whether evening chronotype BD would show differential structural variation in neuroanatomic regions involved in circadian regulation, which might be a proxy for responsiveness to BLT. The anterior inferior hypothalamus contains the SCN and plays a key role in integrating photic and nonphotic inputs that govern circadian rhythms. Based on this rationale, we tested whether hypothalamic structure differs by chronotype in BD. To assess whether such chronotypal differences are specific to BD rather than reflecting a general feature of psychiatric illness, we repeated this comparison for individuals with MDD, psychotic disorders (PY), and healthy controls (HC). To further assess anatomical specificity, we examined for all groups whether any chronotype-related effects were confined to the anterior-inferior hypothalamic region that includes the suprachiasmatic area, or whether similar effects were present across other hypothalamic subunits or the hypothalamus as a whole.

## Methods and Materials

### Subjects

#### UK Biobank

The present study investigated data from the UK Biobank (UKB) (36). Ethical approval for UKB was granted by the North West Multi-centre Research Ethics Committee and was last renewed in 2021, which is valid for five years (Reference: 21/NW/0157, https://www.ukbiobank.ac.uk/about-us/how-we-work/ethics/). Our study was conducted under the UKB Application Number 102266. All participants provided written informed consent for their data to be used for health-related research.

#### Patient selection

Of the 90,088 UKB participants with a baseline structural T1w MRI scan (available at the time of the present study), we selected all available BD patients based on the ICD diagnosis F31x (**Fig. 1**), which resulted in 113 BD patients. These ICD diagnoses were based on linked hospital admission records. To test the specificity of our hypothesis, we compared BD patients against two other major psychiatric groups: MDD and PY. No additional co-morbid psychiatric diagnoses were present in the MDD, BD, or PY groups, except for 15 cases with both BD and PY, which were hierarchically classified as PY to prioritize the more severe diagnosis.. PY patients were selected filtering for an F2x ICD diagnosis, resulting in *N* = 91 patients. MDD patients were pre-selected filtering for an F32x ICD diagnosis, resulting in *N* = 2,595 patients. To make the MDD group size more comparable to the remaining groups, we applied additional filters which also contributed to homogenization of the MDD group. The filters included: exclusion of any psychiatric non-MDD co-morbidities (namely, ICD codes “F0x–2x”, “F30”, “F31”, “F4x–F9x”) or any neurological co-morbidities (namely, ICD codes “Gx”). This reduced the MDD group size to *N* = 642. Next, we further stratified the MDD group to include only those with antidepressant medication (at this point of filtering, the following antidepressants were observed: amitriptyline, escitalopram, citalopram, clomipramine, duloxetine, lofepramine, sertraline, mirtazapine, paroxetine, trazodone, venlafaxine). This reduced the MDD group size to *N* = 205, which we accepted as sufficiently similar as compared to the other groups. Lastly, we selected a group of HC based on having no ICD diagnosis, which resulted in *N* = 9,798 subjects. To achieve comparable group sizes, we randomly subsampled *N* = 200 from the larger pool using Matlab’s “randi” function for uniform random integer selection. The procedure was repeated unitl the subsample matched the BD group in mean age and sex distribution. One HC had was retrospectively excluded due to preprocessing failure (likely caused by corrupted MR data). Demographic and clinical variables, including age, sex, handedness, education, and current mood-stabilizing medication with lithium (given its discussed effects on chronobiology, (37,38)) were further extracted from the UKB for each participant to serve as confounding variables for subsequent statistical testing. One-way analyses of variance (ANOVAs) were utilized to compare age and years of education across the three patient groups and HC, and Pearson’s Chi-squared tests to explore differences in the distributions of biological sex and handedness. We followed-up significant results in the Chi-squared tests with post-hoc testing using standardized residuals to identify the varying pairs in terms of over-/underrepresentation. For post-hoc testing, we assumed a significance threshold of ∣ z ∣>1.96, which roughly corresponds to *p* < 0.05. We visualized significant differences in demographics where applicable using stack bars within RStudio (version 4.2.2, (39)).

**Figure 1.**
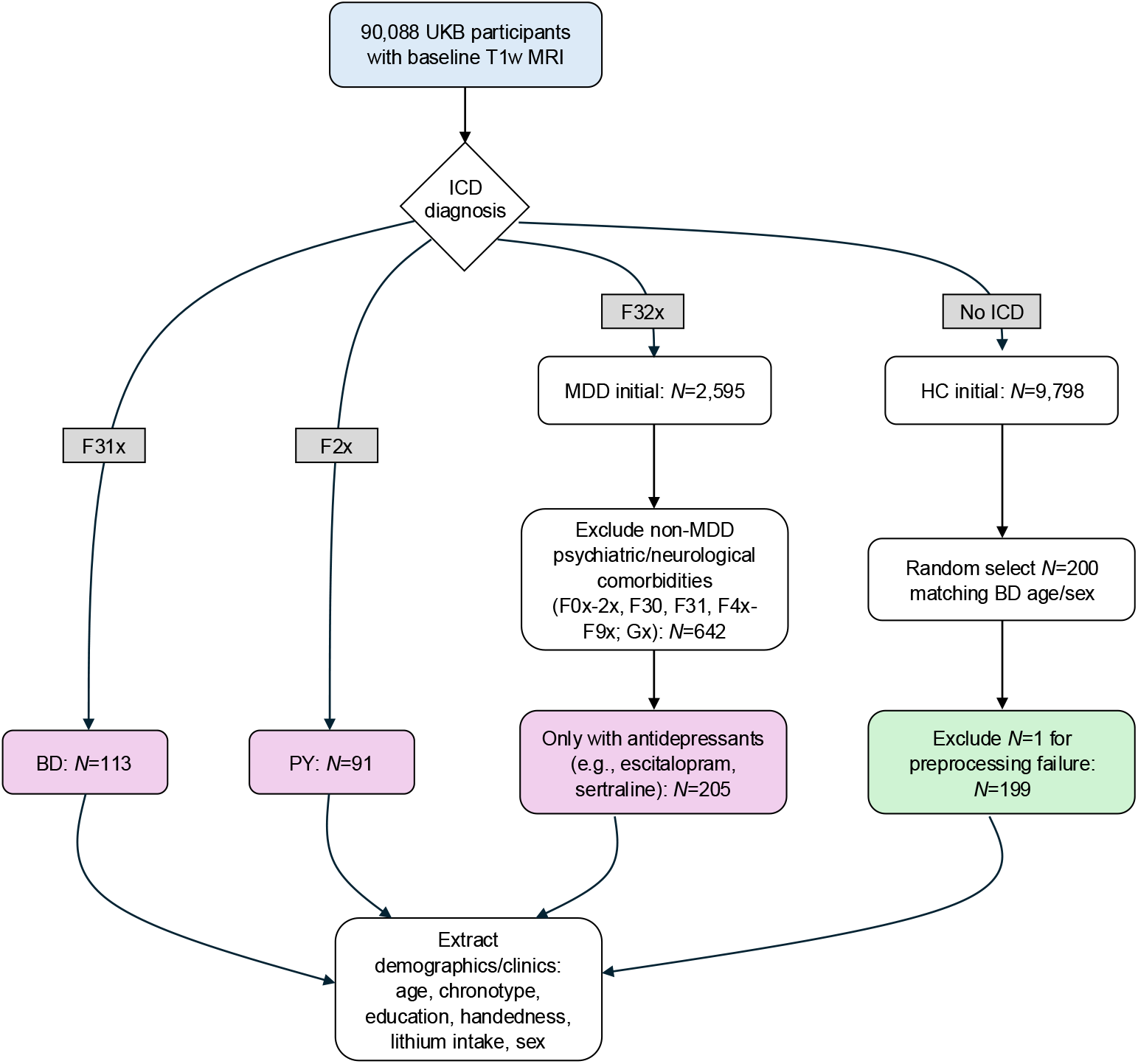
Flow chart showing the data selection process of the present study. Abbreviations: BD = bipolar disorder, HC = healthy controls, ICD = International Classification of Diseases, MDD = major depressive disorder, MRI = magnetic resonance imaging, PY = psychotic disorders, T1w = T1-weighted, UKB = UK Biobank

### Assessment of the chronotype

The chronotype was measured using the self-assessment rating provided within the UKB standard assessment. Participants were asked to rate their own chronotype as one of the following six options: (1) “Definitely a ‘morning’ person”, (2), “More a ‘morning’ than an ‘evening’ person”, (3) “More an ‘evening’ than a ‘morning’ person”, (4) “Definitely an ‘evening’ person”, (5) “Do not know”, and (6) “Prefer not to answer”. These data are provided in the UKB with the field identifier 1180 (using data encoding 100342). We further simplified the chronotypes as either “morning” (collapsing across categories (1) and (2)), “evening” (collapsing across categories (3) and (4)), or “other” (collapsing across categories (5) and (6)). Further details are provided in **Table 1**.

**Table 1.** Demographic details of the study population.

| | Study group | | | | ANOVA [$F$]<sup>+</sup> /<br>Chi-square<br>[$\chi^2$]<sup>++</sup> /<br>Post-hoc SDR<sup>+++</sup> |
| --- | --- | --- | --- | --- | --- |
|  | BD | MDD | PY | HC |  |
| Number of subjects | 113 | 205 | 91 | 199 | n.a. |
| Age<br>[y, mean ± SD] | 54.4 ± 7.64 | 54.1 ± 7.31 | 53.1 ± 7.59 | 53.6 ± 7.29 | $F(3,604) = .682$,<br>$p = .564$ |
| Chronotype<br>[MO/EV/OT] | 63/41/9 | 91/93/21 | 35/39/17 | 103/69/27 | $\chi^2(6) = 13.5$,<br>$p = .036^*$,<br>$z(\text{PY} \times \text{OT}) = 2.06^*$ |
| Education<br>score [mean ± SD] | 13.6 ± 17.1 | 12.5 ± 13.9 | 14.6 ± 13.7 | 11.4 ± 11.9 | $F(3,545) = 1.16$,<br>$p = .325$ |
| Handedness<br>[R/L/A] | 96/14/3 | 182/19/4 | 84/4/3 | 183/15/1 | $\chi^2(6) = 8.09$,<br>$p < .232$ |
| Lithium intake<br>[yes/no] | 21/92 | 1/204 | 2/89 | 0/199 | $\chi^2(3) = 79.2$,<br>$p < .001^*$ |
| Sex<br>[F/M] | 61/52 | 146/59 | 44/47 | 98/101 | $\chi^2(3) = 25.0$,<br>$p < .001^*$,<br>$z(\text{MDD} \times \text{F}) = 4.91^*$,<br>$z(\text{HC} \times \text{M}) = 2.84^*$ |

**Abbreviations:** ANOVA = analysis of variance, A = ambidextrous, BD = bipolar disorder, EV = evening chronotype, F = female, HC = healthy control, L = left-handed, M = male, MDD = major depressive disorder, MO = morning chronotype, N = sample size, n.a. = not applicable, OT = unclear chronotype (“other”), PY = psychotic patients, R = right-handed, SD = standard deviation, SDR = standardized residuals, y = years
**Symbols:** <sup>+</sup> = ANOVAs were performed to test differences of age and years of education between the four study groups; <sup>++</sup> = Pearson’s Chi-squared tests were performed to test differences of sex, handedness, lithium intake, and chronotype frequencies between the four study groups; <sup>+++</sup> = for significant $\chi^2$ tests (except for lithium intake, since hardly present in non-BD), we investigated standardized residuals post-hoc to determine the differing group(s), assuming a significance threshold of $|z| > 1.96 \approx p < 0.05$ ; \* = significant at an alpha-level of $p \leq .05$

### Neuroimaging data acquisition

All UKB subjects underwent a standardized brain imaging protocol on Siemens Skyra 3T systems with a standard Siemens 32-channel RF receiver head coil. The present study only considered baseline scans, acquired since 2014. Structural T1-weighted (T1w) MRI data were acquired as sagittal slices using a 3D MPRAGE sequence with the following image parameters: field of view = 256×256×208 matrix, voxel resolution = 1 mm isotropic, iPAT in-plane acceleration factor = 2, prescan normalization = true, total scan duration = 5 minutes. The full details of UKB neuroimaging data acquisition can be found here: https://biobank.ctsu.ox.ac.uk/crystal/crystal/docs/brain_mri.pdf.

### Neuroimaging data analysis

The primary goal of the present study was to contrast morphological alterations of distinct hypothalamic subunits as they interact with the chronotype in BD as compared to other major psychiatric disorders. We defined morphological alterations as differences in volume. The software FreeSurfer (40) offers a fully-automated pipeline to segment five different hypothalamic subunits, separately for left and right hemispheres, and calculate the respective volumes (41). The five hypothalamic subunits include: (1) **anterior-inferior**, containing the suprachiasmatic and the supraoptic nuclei, (2) **anterior-superior**, containing the preoptic area and parts of the paraventricular nucleus, (3) **posterior**, containing the mamillary bodies, parts of the lateral hypothalamus and the tuberomamillary nucleus, (4) **inferior tubular**, containing the infundibular (or arcuate) and the ventromedial nuclei as well as the lateral tubular tubulus, and (5) **superior tubular**, containing the dorsomedial nucleus, parts of the paraventricular nucleus, and parts of the lateral hypothalamus. The algorithm uses a convolutional neural network and was trained by applying aggressive data augmentation, which increases robustness against varying sequence parameters and anatomy (such as often observed in disease). To note, to run the hypothalamic segmentation pipeline, T1w data first need to be processed with FreeSurfer’s automated processing pipeline for structural data (“recon-all”). We completed this using FreeSurfer version 7.4.1. In brief, recon-all performs cortical surface reconstructions, volumetric segmentations and parcellations. Preprocessing of T1w data includes motion/intensity correction, skull stripping, registration, subcortical labeling, topological fixes, white/pial surface modeling, and atlas-based parcellation. The details of this pipeline are described in (40,42,43) or online at the FreeSurfer Wiki (https://surfer.nmr.mgh.harvard.edu/fswiki/recon-all).

### Statistics

To investigate an interaction effect between diagnosis and chronotype with respect to the volume of the SCN-containing hypothalamic subunit, namely the anterior-inferior subunit, we employed a one-way ANOVA. In the ANOVA, we corrected for confounding effects of age, handedness, current lithium medication (binarized as “yes” or “no”), and sex. Notice that the UKB provides data on the level of education (“Education score England”, field ID 26414) which estimates area-level deprivation in education (see the UKB documentation for further details at https://biobank.ndph.ox.ac.uk/ukb/field.cgi?id=26414). However, 59 out of the total of our 608 participants did not have available education scores, such that considering education as an additional confounding variable would have decreased the power of our study. We therefore chose to exclude education as confounder from our model. Descriptive statistics of the education score variable are however provided in **Table 1**. To test the specificity of a potential interaction to the SCN-containing hypothalamic subunit, we also investigated interaction effects between diagnosis and chronotype with respect to the volumes of the four remaining segmented hypothalamic subunits (anterior-superior, posterior, inferior-tubular, superior-tubular), as well as for the entire hypothalamic volume. All subunits were collapsed across the two hemispheres, normalized by the total intracranial B) volume, and analyzed using analog ANOVAs. We further investigated significant interaction effects in post-hoc estimated marginal means (EMM) testing of pairwise comparisons. All statistics were calculated within *RStudio* (version 4.2.2, (39)). EMM was carried out within the *RStudio* package “emmeans” (10.32614/CRAN.package.emmeans, (44)). For the omnibus ANOVA tests, we assumed a Bonferroni-corrected alpha-level of *p* <.01 (correcting for five separate tests, one for each subunit). EMM results were family-wise error (FWER) corrected using Tukey’s method for comparing a family of three estimates.

## Results

### Subjects

After applying the subject selection criteria as specified above (cf. also **Fig. 1**), data from 608 UKB participants were included into our study. Of those, 113 had were diagnosed with BD, 205 with MDD, 91 with PY, along with 199 HC. One-way ANOVAs suggested no differences in age means [*F*(3,604) =.682, *p* =.564] and mean education scores [*F*(3,545) = 1.16, *p* =.325] across the four groups. Chi-squared tests suggested no difference in the distribution frequencies of handedness [*χ*^2^(6) = 8.09, *p* <.232]. As expected, Chi-squared tests indicated a significant difference in the frequency of current lithium intake [*χ*^2^(3) = 79.2, *p* <.001], since hardly all patients with prescribed lithium medication were in the BD group [*N*_*BD*_ = 21, *N*_*MDD*_ = 1, *N*_*PY*_ = 2, *N*_*HC*_ = 0].

Moreover, Chi-squared tests indicated a significant difference in the frequency of chronotype distributions [χ^2^(6) = 13.5, *p* =.036]. Post-hoc EMMs identified more “Other” (i.e., unclear) chronotypes in the PY group than expected as compared to the other three groups [*z*(PYxOT) = 2.06, *p* <.05, Fig. 2A]. Lastly, Chi-squared tests indicated a significant difference in the frequency of sex distributions [*χ*^2^(3) = 25.0, *p* <.001]. Post-hoc EMMs identified more females in the MDD group than expected as compared to the other three groups [z(MDDxF) = 4.91, p <.05, **Fig. 2B**]] and more males in the HC group than expected as compared to the other three groups [z(HCxM) = 2.84, p <.05, **Fig. 2B**]. Further statistical details on subject demographics are provided in **Table 1**.

**Figure 2.**
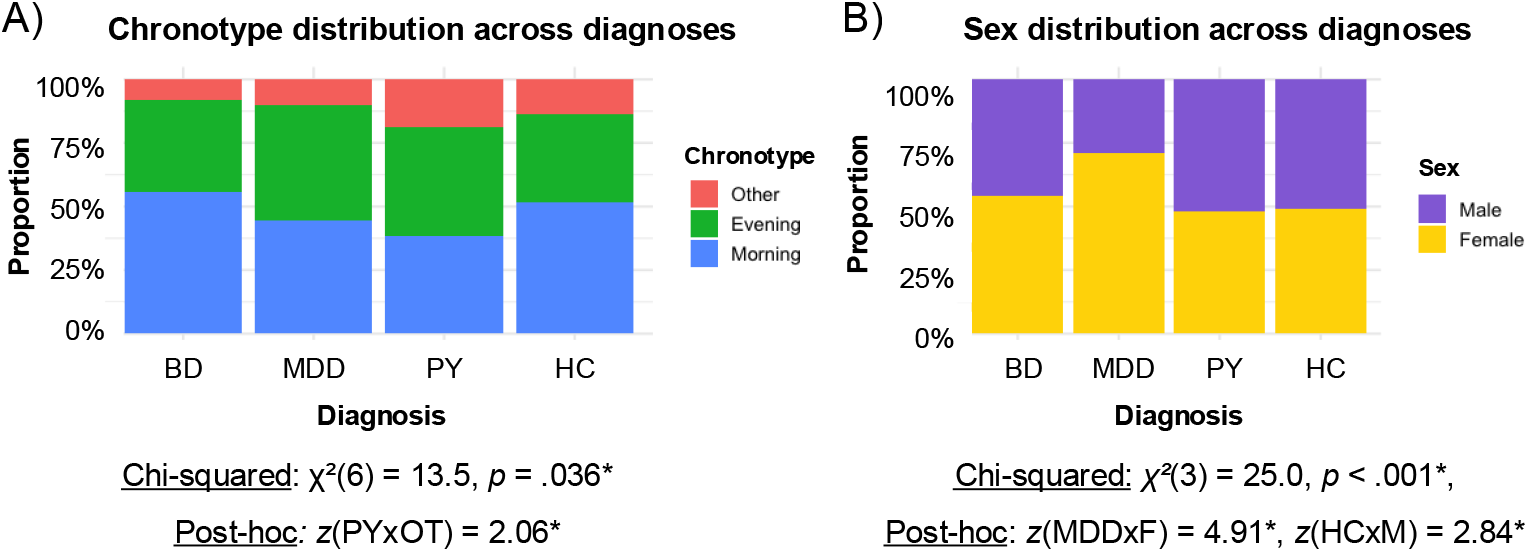
Demographic differences across the study groups. Differences in chronotype (A) and sex (B) distributions were observed across diagnostic groups (see Table 1). Chronotype category “Other” was more frequent in the psychotic disorders group, while sex distribution differed primarily between the major depressive disorder and healthy control groups. All demographic variables shown here were included as covariates in the main analyses. Abbreviations: ANOVA = analysis of variance, EMM = estimated marginal means, F = female, HC = healthy controls, M = male, MDD = major depressive disorder, OT = “other” chronotype (referring to unclear participant self-assessment), PY = psychotic disorders

### Segmentation results

The automated hypothalamic subunit segmentation succeed in all but one case (1 HC, which was excluded from subsequent analyses, cf. also **Fig. 1**). Visual inspection of the results suggested valid segmentation results based on known macroscopic neuroanatomy of the whole hypothalamus (**Fig. 3A**) and especially of the anterior-inferior unit (**Fig. 3B**). The anterior-inferior subunit contains the SCN (along with the supraoptic nucleus), and was therefore the main region-of-interest in our study. As shown in **Figure 3B** on the right, the SCN-containing region was accurately located superior-posterior to the optic chiasm and superior to the optic tract, which matches the known neuroanatomical ground truth (45). Moreover, the segmentation suggested slight morphological differences in bilateral subunits, which matches the widespread left-right asymmetry found in the human brain (46).

**Figure 3.**
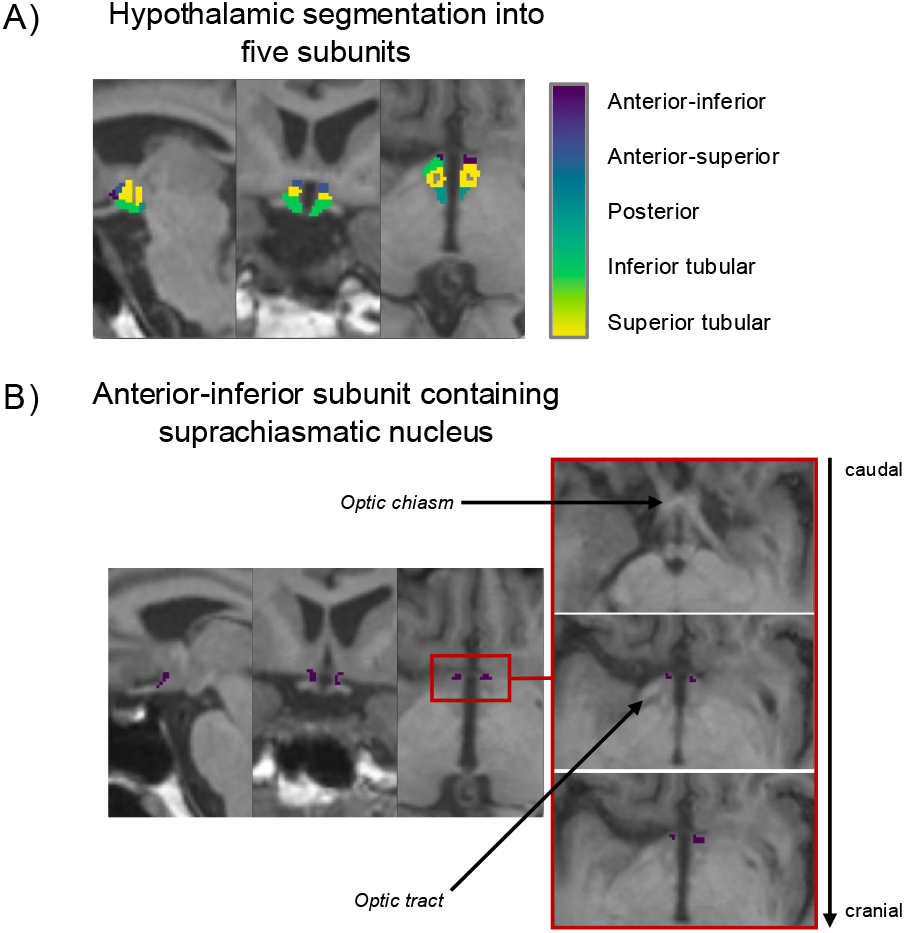
Segmentation results for the five assessed hypothalamic subunits. We used a fully-automated AI-trained segmentation algorithm, provided within FreeSurfer, to segment five subunits of the hypothalamus, for each subject individually. We here provide exemplary segmentation results for one healthy subject of our study population. In (A), we provide an overview of all five subunits (notice that for the present study, we collapsed across the hemispheres since we had no hemispheric-specific hypothesis, although the default FreeSurfer output provides each subunit as separate labels for right and left hemispheres). In (B), we zoom in on the segmentation of the anterior-inferior segment, since it contains the suprachiasmatic nucleus (SCN, next to the supraopticus nucleus). The suprachiasmatic nucleus was the main region-of-interest in the present study. To demonstrate the validity of our segmentation results, we provide a separate panel in B (right), in which we provide a row of ascending axial slices, starting caudally at the optic chiasm, separated by ∼1mm each. These slices illustrate the anatomical location of the segmented anterior-inferior hypothalamic subunit relative to the optic chiasm and optic tract. Notice also the differences in the segmentation across the hemispheres.

### Interaction effects between diagnosis and chronotype

One-way ANOVAs were used to test for an interaction between diagnosis and chronotype with respect to the volume of the SCN-containing anterior-inferior hypothalamic subunit (**Fig. 4A**) and the four remaining hypothalamic subunits, to explore specificity (**Fig. 4B–E**). The analog results for the collapsed hypothalamic volume are provided in **Figure 4F**. ANOVAs were corrected for age, handedness, current lithium intake, and sex. After multiple comparisons correction, the interaction between diagnosis and chronotype was significant only for the volume of the anterior-inferior subunit (*F*(6, 590) = 2.87, *p* =.009). The full statistical details for all subunits are provided in **Table 2**. To identify the varying pairs, the significant ANOVA interaction effect between diagnosis and chronotype for the anterior-inferior subunit was followed up with post-hoc EMM testing (**Table 3**, **Fig. 4A**). EMM revealed a significantly higher volume of that subunit in “Evening” vs. “Morning” chronotypes specifically in BD (*t* = 3.24, *p*_*FWER*_ =.004). Moreover, we found a significantly higher volume in “Morning” vs. “Other” (i.e., unclear) chronotypes in PY, which was however a weaker effect as compared to the effect found in BD_Evening_ vs. BD_Morning_ (*t* = 2.81, *p*_*FWER*_ =.014).

**Table 2.** Statistical assessment of an interaction effect between diagnosis and chronotype with respect to different hypothalamic subunits (and the whole hypothalamic volume). One-way a ANOVAs were used to test for an interaction, corrected for age, handedness, current lithium intake, and sex. We assumed a Bonferroni-corrected alpha level of *p* ≤.01 indicating significance (accounting for the five separately tested hypothalamic subunits). Significant interactions were followed-up in post-hoc testing (cf. Table 3 and main text).

|  | Anterior-inferior subunit |  |  |  |  | Anterior-superior subunit |  |  |  |  |
| --- | --- | --- | --- | --- | --- | --- | --- | --- | --- | --- |
|  | DOF | Sum Square | Mean Square | F-value | p-value | DOF | Sum Square | Mean Square | F-value | p-value |
| Diagnosis | 3 | 6.5e-11 | 2.2e-11 | .562 | .640 | 3 | 1.5e-10 | 5.1e-11 | 1.22 | .302 |
| Age | 1 | 6.4e-11 | 6.4e-11 | 1.67 | .197 | 1 | 4.3e-10 | 4.3e-10 | 1.3 | .001** |
| Chronotype | 2 | 6.100e-11 | 3.068e-11 | .802 | .449 | 2 | 1.1e-10 | 5.7e-11 | 1.38 | .252 |
| Handedness | 2 | 6.9e-11 | 3.5e-11 | .905 | .405 | 2 | 7.0e-11 | 3.5e-11 | .843 | .431 |
| Lithium intake | 1 | .0e-00 | 3.0e-14 | .001 | .978 | 1 | 2.0e-12 | 2.1e-12 | .050 | .824 |
| Sex | 1 | 1.0e-10 | 1.0e-10 | 2.68 | .102 | 1 | 2.9e-10 | 2.9e-10 | 7.00 | .008** |
| Diagnosis x Chronotype | 6 | **6.6e-10** | **1.1e-10** | **2.87** | **.009\*\*** | 6 | 3.6e-10 | 5.9e-11 | 1.43 | .199 |
| Residuals | 590 | 2.3e-08 | 3.8e-11 | n.a. | n.a. | 590 | 2.5e-08 | 4.2e-11 | n.a. | n.a. |
|  | Posterior subunit |  |  |  |  | Inferior tubular subunit |  |  |  |  |
|  | DOF | Sum Square | Mean Square | F-value | p-value | DOF | Sum Square | Mean Square | F-value | p-value |
| Diagnosis | 3 | 4.0e-09 | 1.3e-09 | 2.46 | .062 | 3 | 5.0e-09 | 1.7e-09 | 2.82 | .038* |
| Age | 1 | 1.3e-09 | 1.3e-09 | 2.33 | .127 | 1 | 4.8e-09 | 4.8e-09 | 8.05 | .005** |
| Chronotype | 2 | 3.1e-09 | 1.6e-09 | 2.90 | .056 | 2 | 7.0e-10 | 3.3e-10 | .556 | .574 |
| Handedness | 2 | 7.0e-10 | 3.7e-10 | .685 | .505 | 2 | 8.0e-10 | 4.0e-10 | .672 | .511 |
| Lithium intake | 1 | 3.0e-10 | 2.9e-10 | .535 | .465 | 1 | 2.2e-09 | 2.2e-09 | 3.66 | .056 |
| Sex | 1 | 4.0e-09 | 4.9e-09 | 7.43 | .007** | 1 | 3.0e-10 | 3.1e-10 | .516 | .473 |
| Diagnosis x Chronotype | 6 | 2.6e-09 | 4.4e-10 | .813 | .560 | 6 | 5.1e-09 | 8.6e-10 | 1.44 | .196 |
| Residuals | 590 | 3.2e-07 | 5.4e-10 | n.a. | n.a. | 590 | 3.5e-07 | 6.0e-10 | n.a. | n.a. |
|  | Superior tubular subunit |  |  |  |  | [Whole hypothalamic volume] |  |  |  |  |
|  | DOF | Sum Square | Mean Square | F-value | p-value | DOF | Sum Square | Mean Square | F-value | p-value |
| Diagnosis | 3 | 5.6e-10 | 1.9e-10 | .648 | .584 | 3 | 1.9e-08 | 6.5e-09 | 1.99 | .114 |
| Age | 1 | 7.e-10 | 7.0e-10 | 2.44 | .119 | 1 | .0e00 | 3.0e-11 | .009 | .923 |
| Chronotype | 2 | 2.3e-10 | 1.2e-10 | .401 | .670 | 2 | 6.0e-09 | 3.0e-09 | .919 | .400 |
| Handedness | 2 | 3.5e-10 | 1.7e-10 | .601 | .549 | 2 | 5.6e-09 | 2.8e-09 | .861 | .423 |
| Lithium intake | 1 | 2.9e-10 | 2.9e-10 | 1.01 | .317 | 1 | 2.0e-09 | 2.0e-09 | .625 | .430 |
| Sex | 1 | 5.4e-10 | 5.4e-10 | 1.88 | .171 | 1 | 1.7e-08 | 1.7e-08 | 5.31 | .022 |
| Diagnosis x Chronotype | 6 | 1.6e-09 | 2.7e-10 | .936 | .468 | 6 | 2.4e-08 | 3.9e-09 | 1.21 | .298 |
| Residuals | 590 | 1.7e-07 | 2.9e-10 | n.a. | n.a. | 590 | 1.9e-06 | 3.2e-09 | n.a. | n.a. |

Abbreviations: ANOVA = analysis of variance, DOF = degrees of freedom, n.a. = not applicable
Symbols: \* = significant at an uncorrected alpha level of $p \leq .05$ ; \*\* = significant at a Bonferroni-corrected alpha level of $p \leq .01$ ; results surviving the main interaction effect of interest are printed in **bold**

**Table 3.**
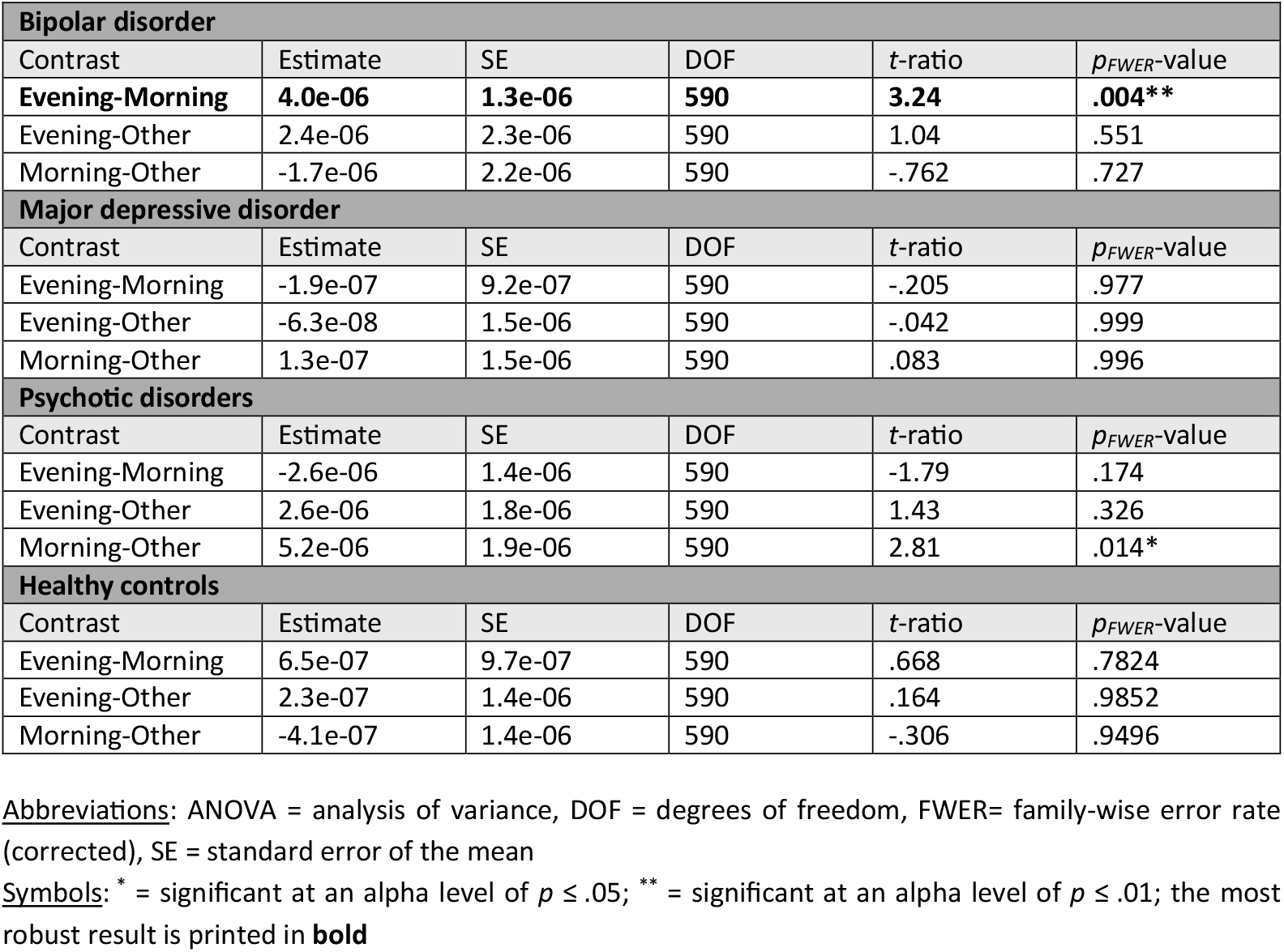
Post-hoc assessment of significant ANOVA interaction effects (diagnosis x chronotype). One way ANOVA testing indicated a significant interaction effect for diagnosis x chronotype specifically for the anterior-inferior hypothalamic subunit (cf. Table 2 and main text). We explored this effect further in post-hoc testing to identify the varying pairs. We used estimated marginal means testing, averaged over the levels of handedness, current lithium intake, and sex, and family-wise error-corrected using Tukey’s method for comparing a family of three estimates.

**Figure 4.**
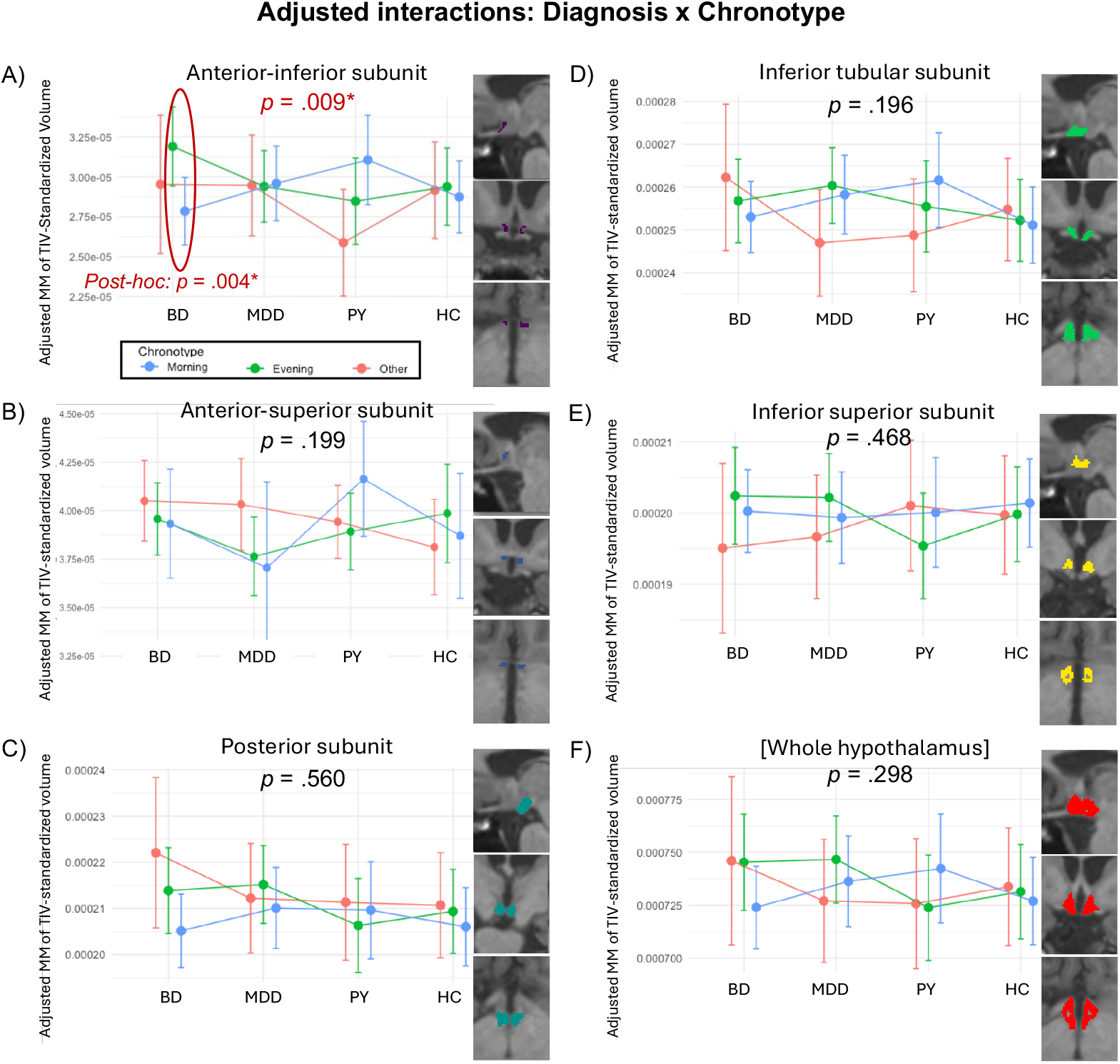
Chronotype related differences in hypothalamic subunit volumes across diagnostic groups. Panels A-E show results for individual hypothalamic subunits, and panel F shows total hypothalamic volume. A diagnosis by chronotype interaction was observed specifically for the anterior inferior hypothalamic subunit (A), with larger volumes in evening compared with morning chronotypes in bipolar disorder. No comparable effects were observed for other subunits or for total hypothalamic volume. Statistical models were adjusted for age, sex, handedness, lithium use, and total intracranial volume (see Table 2 and Table 3 for details). Abbreviations: ANOVA = analysis of variance, BD = bipolar disorder, (E)MM = (estimated) marginal means, HC = healthy controls, MDD = major depressive disorder, PY = psychotic disorders, TIV = total intracranial volume, * = significant at a corrected alpha-level of p ≤.01.

## Discussion

The present study examined whether hypothalamic structure differs by chronotype in bipolar disorder, motivated by prior evidence that evening chronotype may index more severe clinical features and poorer treatment response (29). We identified a diagnosis-specific association between chronotype and hypothalamic morphology, localized to an anterior inferior hypothalamic subunit encompassing the suprachiasmatic region. Individuals with BD and an evening chronotype showed larger volumes of this subunit compared with those who had morning chronotypes, an effect that was not observed in individuals with MDD, PY, or controls. Importanty, no comparable chronotype related effects were detected in other hypothalamic subunits or in total hypothalamic volume. Together, these findings indicate that chronotype related structural variation in circadian relevant hypothalamic anatomy is detectable in BD and shows both diagnostic and anatomical specificity.

One possible interpretation of these findings is that chronotype captures meaningful variation in circadian system organization in BD. The anterior inferior hypothalamus contains the suprachiasmatic nucleus, the central circadian pacemaker, and plays a key role in integrating photic and nonphotic inputs that regulate circadian rhythms (41). Structural variation in this region may therefore reflect long term differences in circadian signaling, regulation, or adaptation. While evening chronotype represents a normative variation in circadian preference (47), its interaction with bipolar pathology may be associated with altered circadian system organization that is not observed in other psychiatric conditions. In this sense, chronotype may function as a modifier of circadian biology in BD rather than as an independent pathological factor (5,48).

The observation that chronotype related effects were only observed for bipolar disorder warrants consideration in light of proposed differences in circadian involvement across psychiatric diagnoses (49,50). Although circadian dysregulation has been described in multiple psychiatric conditions, it has been argued to be particularly prominent in BD, both clinically and biologically (51). Within this framework, chronotype may differentially interact with disorder specific pathophysiological processes, resulting in detectable neuroanatomical differences in circadian relevant regions in BD but not in other diagnostic groups. However, such interpretations remain preliminary and should be understood as hypotheses rather than conclusions drawn from the present data.

Alternative explanations for the observed volumetric differences must also be considered. The anterior-inferior hypothalamic subunit analyzed here encompasses the suprachiasmatic nucleus as well as adjacent nuclei, including the supraoptic nucleus, which plays an important role in neuroendocrine regulation (41). Accordingly, the observed structural variation likely reflects properties of this broader circadian relevant hypothalamic region rather than the SCN in isolation. In addition, volumetric differences measured with structural MRI may arise from multiple underlying cellular or microstructural processes, such as variation in neuronal or glial composition, vascular factors, or developmental influences, which cannot be disentangled with the present methodology (52,53). Finally, given the cross-sectional design, the direction of causality remains unresolved. Importantly, the effect is specific to the diagnosis × chronotype interaction in BD, rather than a general chronotype effect. Thus, the volumetric differences likely reflect unique processes arising from the combination of BD pathology and evening chronotype, potentially involving bidirectional or adaptive mechanisms, rather than intrinsic circadian pacemaker properties per se (54,55).

Several limitations merit comment. Because the study is cross sectional, the present findings do not resolve the temporal ordering of chronotype related differences in hypothalamic structure. Such differences could precede the onset of bipolar disorder, emerge in association with illness related circadian disruption, or reflect adaptive or maladaptive changes over the course of the disorder (50,56). Chronotype was assessed using self report measures, which capture stable circadian preference but do not directly index physiological circadian phase (30,57). Another limitation of this study is that we controlled only for lithium use and not for other psychotropic medications, which may also influence brain structure, such that their potential effects cannot be fully excluded. Future larger studies with comprehensive medication covariate adjustment are needed to confirm the specificity of our findings. In addition, the UK Biobank sample is predominantly middle aged to older adults (36), and future work will be needed to determine whether similar associations are present earlier in the course of illness.

These considerations point to several promising directions for further research. Longitudinal studies tracking hypothalamic structure alongside circadian function and clinical course will be well suited to clarify the stability and temporal dynamics of chronotype related structural differences (58). Integrating structural imaging with physiological measures of circadian timing, such as melatonin onset or actigraphy, may help link hypothalamic morphology more directly to circadian function (59). Moreover, examining afferent pathways to circadian hypothalamic regions, including retinal structure and retinohypothalamic tract integrity (45), could provide additional insight into whether altered sensory input contributes to structural variation in circadian related neuroanatomy (60).

From a clinical perspective, these findinigs further highlight chronotype as a biologically meaningful feature of BD pathophysiology (61–63). The observed association between chronotype and circadian relevant hypothalamic structure provides neuroanatomical support for the broader view that circadian related factors contribute to BD outcomes and that chronotype captures differences in circadian system organization (6,14). While the present results do not by themselves establish utility for individual level prediction or treatment stratification, they motivate additional investigation into whether hypothalamic morphology relates to circadian dysfunction, treatment response, or clinical outcomes (64,65). Such work will be important to further determine whether and how chronotype informed approaches can contribute to more personalized circadian based interventions in bipolar disorder (66).

## Data Availability

The data used in this study were obtained from the UK Biobank under approved application access. UK Biobank is a large scale biomedical database that provides controlled access to its data for qualified researchers subject to application and approval. The data are not publicly available but can be accessed through the UK Biobank repository by researchers who meet the access criteria. The analysis scripts used in this study are available from the corresponding author upon reasonable request.

https://ams.ukbiobank.ac.uk/ams

## CRediT authorship contribution statement

**MT:** Conceptualization, Data curation, Formal analysis, Methodology, Visualization, Writing – original draft, Writing – review & editing. **JR:** Formal analysis, Validation, Writing – review & editing. **ES:** Funding acquisition, Writing – review & editing. **DS:** Conceptualization, Writing – review & editing. **PH:** Conceptualization, Funding acquisition, Supervision, Writing – review & editing.

## Acknowledgment

MT is supported by a project grant from the Hans und Marianne Schwyn-Stiftung and by a Filling-the-Gap fellowship of the Faculty of Medicine, University of Zurich.

